# Repurposing Colchicine for the Management of COVID-19: A Systematic Review and Meta-analysis

**DOI:** 10.1101/2021.02.17.21251884

**Authors:** Rashmi Ranjan Mohanty, Bikash Ranjan Meher, Biswa Mohan Padhy, Smita Das

## Abstract

Many anti-inflammatory drugs like, tocilizumab, N-acetylcystiene and etolizumab has been repurposed for the management of COVID-19 with variable success. Colchicine exhibits anti-inflammatory activity by tubulin disruption and inhibition of leucocyte-mediated inflammatory activities like production of superoxide and release of various cytokines which are central to the pathophysiology of COVID-19. So, this systematic review and meta-analysis assessed the currently available data on the use of colchicine for the treatment of COVIDLJ19. A total of 3 studies (2 RCTS and 1 observational study) including 402 patients were included out of which 194 patients received colchicine. The random effect model showed the overall pooled OR to be 0.32 (95%CI: 0.18 to 0.56) for the primary outcome (Clinical deterioration) which was statistically significant (p <0.0001). Also there was increase in adverse effect (diarrhoea) with use of colchicine as suggested by the pooled OR (OR=4.56, 95%CI: 2.04 to 10.15, P=0.0002). With the limited number of available studies, it has shown statistically significant reduction in clinical deterioration in COVID-19. Though there was an increase in adverse effect in the form of diarrhoea, it was mild and self-limiting.

## Introduction

Since its inception in November 2019, Severe acute respiratory syndrome coronavirus 2 (SARS-CoV-2) has become a pandemic affecting more than 195 countries with approximately 31 million infections and 0.97 million deaths till the end of September 2020 [1].Though viral pneumonia is the common clinical presentation, the major complication of coronavirus disease 2019 (COVID-19) is due to immune activation, hyper inflammatory state, prothrombotic state, and cytokine storm [2,3]. All these pathologic processes leads to acute respiratory distress syndrome (ARDS), which is a life-threatening form of acute lung injury [4]. The inflammatory mediators responsible for cytokine storm and ARDS, like interleukin 1 (IL1), interleukin 6 (IL6), and tumor necrosis factors (TNFs) have been found to increase in severe COVID-19 [5]. Therefore, many anti-inflammatory drugs like tocilizumab, N-acetylcystiene, and etolizumab have been repurposed for management of COVID-19 with variable success [6-8].

Colchicine is a lipid-soluble alkaloid derived from *Colchicum autumnale* [9]. Currently it is being used for many auto-inflammatory conditions like gout, Behcet’s disease, and familial Mediterranean fever [10]. Tubulin disruption and subsequent down regulation of inflammatory pathway is the major mechanism of action of colchicine [11]. An *in vitro* study has demonstrated that the internalization of the coronavirus is dependent on tubulin function [12]. Also colchicine inhibits leucocyte-mediated inflammatory activities like the production of superoxide and release of various cytokines [13]. Based on these anti-inflammatory properties of colchicine, researchers have proposed its use in COVID-19 [14,15]. A case report by Mansouri *et al*, demonstrated the beneficial effect of colchicine in COVID-19 associated cytokine release syndrome [16]. Similarly a case series of 9 patients reported the favourable outcome by colchicine use in COVID-19 [17]. Additionally, few randomized controlled trials (RCTs) and observational studies have also been carried out to assess the clinical efficacy of colchicine in the management of COVID-19 [18-20].

Hence, we conducted this systematic review and meta-analysis to assess the currently available data to rationalize the repurposing of colchicine for the treatment of COVIDLJ19 as add on therapy. This review may provide clinicians an overview of contemporary scientific evidence regarding the use of colchicine in the management of COVIDLJ19 patients.

## Materials and methods

### Development and registration of protocol

The protocol was written in accordance with PRISMA-P guideline and registered in the prospective register of a systematic review (PROSPERO). The PROSPERO registration number is CRD42020209814.

### Types of studies

All RCTs and observational studies comparing the use of colchicine in COVID 19 with usual care or placebo were included for the analysis. Clinical outcome and adverse effects were the outcome measures in all the included studies. The study inclusion was not restricted by year of publication, site of study, and dose of colchicine used. The studies published only in the English language were included.

### Types of Participants

RT-PCR confirmed COVID-19 adult patients of both genders treated with usual care with or without the addition of colchicine were included in the study.

### Types of intervention

The intervention was defined as the administration of colchicine in COVID 19 patients along with usual care irrespective of the dose, timing, or frequency.

### Types of comparator

The comparison was between usual care with colchicine versus usual care alone or with a placebo.

### Outcome measures

#### Primary outcome

1. Clinical deterioration (within the follow up time available in each study).

Clinical deterioration was defined as the need for oxygen supplementation or ICU care or death

#### Secondary outcomes

1. Development of adverse effect

The adverse effect was defined as the development of diarrhoea (as it is the most common adverse effect of colchicine) and/or any other adverse effect leading to the withdrawal of colchicine.

### Information source

PubMed, EMBASE, the Cochrane Library, SCOPUS, and Web of Science were searched for articles on colchicine therapy in COVID 19 from inception till September 15, 2020. The reference lists of retrieved articles were checked for additional studies. For unpublished data, we checked the International Clinical Trials Registry Platform (ICTRP), which is a central database containing trial registration data sets provided by the different international trial registries including ClinicalTrials.gov. Pre-print server medRxiv and bioRxiv were also searched for pre-print data.

### Search strategy

A combination of subject terms and keywords were used and appropriate adjustments of vocabulary and grammar between different databases were done using the PICO method. The search used both Medical Subject Headings (MeSH), as well as keyword variants of all relevant terms. A combination of keywords and Boolean operators like “Colchicine” OR “Anti-inflammatory drug” AND “COVID-19” OR “Severe acute respiratory syndrome coronavirus 2” was used for designing the search algorithm.

### Data extraction and management

Three review authors (RRM, BRM, SD) independently extracted and assessed the quality of data using the predefined eligibility criteria following Cochrane Collaboration’s guidelines. Any disagreement between them was resolved by the fourth author (BMP). A pre-designed data extraction format was used for the recording of data which includes study design, basic information, treatment details, and outcome measures.

### Assessment of risk of bias in included studies

Risk of bias-2 (RoB-2) tool for RCTs and Risk of Bias in Non-Randomized Studies -of Interventions (ROBINS-I) for the observational study were used to assess the risk of bias.

### Data analysis

Cochrane Program Review Manager 5.3 software was used for the Meta-analysis. For dichotomous values, odd’s ratio (OR) and 95% confidence interval was expressed in accordance with Cochrane Handbook for Systematic Reviews of Interventions. I^2^ statistic was used to check heterogeneity among eligible studies. The random-effect model was used for data synthesis.

### Assessment of publication bias

A funnel plot was used to assess the presence of publication bias.

### Grade of evidence

GRADE profiler software (V 3.6.1) was used for quality assessment of the evidence.

## Results

### Description of studies

All database searches resulted in a total of 97 studies describing the use of colchicine in COVID-19. Fifty two studies from PubMed, 22 on-going registered clinical trials, and 23 studies from pre-print server medRxiv were retrieved. After reviewing abstracts and full text, 3 studies (2 RCTs and 1 observational study) were selected for systematic review and meta-analysis as per the inclusion criteria. The PRISMA flowchart of study selection is depicted in Figure 1.

**Figure 1:**
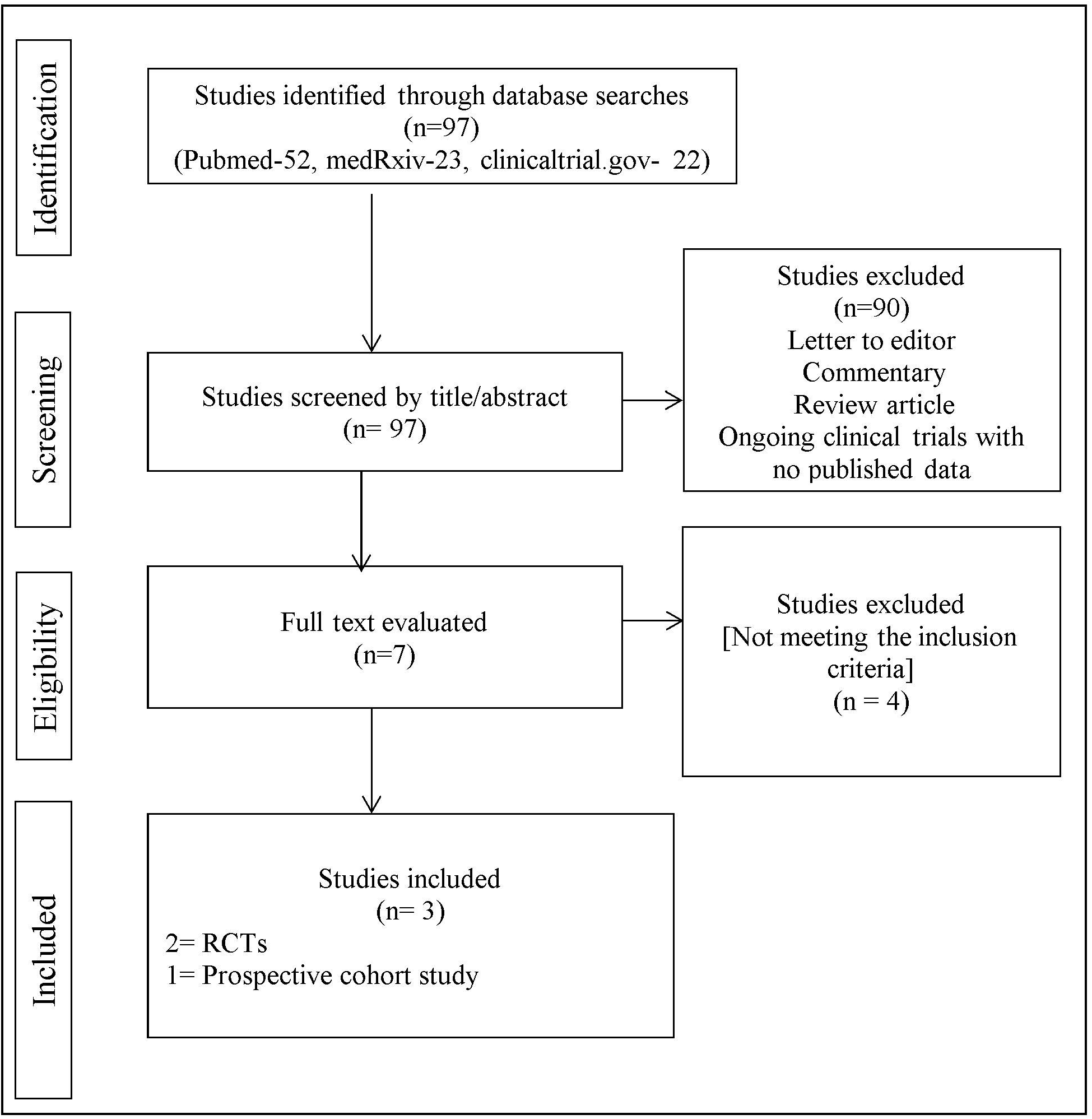
PRISMA flowchart of study selection process

The study by Deftereos *et al*, was a prospective open-label randomized controlled trial and Lopes *et al*, was a randomized, double-blinded, placebo-controlled clinical trial [18,19]. Though the study by Scarsi *et al*, was an observational cohort study, the selection bias can be considered minimal as only patients with contraindication to the drug (ie, with renal failure) were excluded [20]. Similarly, as the interval between hospitalisation and colchicine initiation, was very short (mean of 1 day), the risk of survival bias can also be considered minimal in Scarsi *et al*. The characteristics of included studies are depicted in table 1.

A total of 402 RT-PCR positive patients were included in the 3 studies. Out of them, 194 patients received colchicine along with usual care. In all the included studies, the average age, gender variability, and comorbidities were comparable in both the groups. The average period of follow up ranged from 7 days to 21 days. The dose of colchicine varied from 0.5 milligrams to 1.5 milligrams per day. The duration of treatment with colchicine was 7 days in Lopes *et al*, 10 days in Scarsi *et al* and 21 days in Deftereos *et al*. The characteristics of patients included in the meta-analysis are depicted in table 2.

### Risk of bias in the included studies

The risk of bias assessment for all studies was carried out for the primary outcome. The use of RoB-2 revealed that both the RCTs had “some concerns” regarding the risk of bias. In the observational study included in the meta-analysis, there was a “serious” risk of bias as per ROBINS-I. The result of the risk of bias assessment of RCTs and observational study is depicted in figure 2.

**Figure 2:**
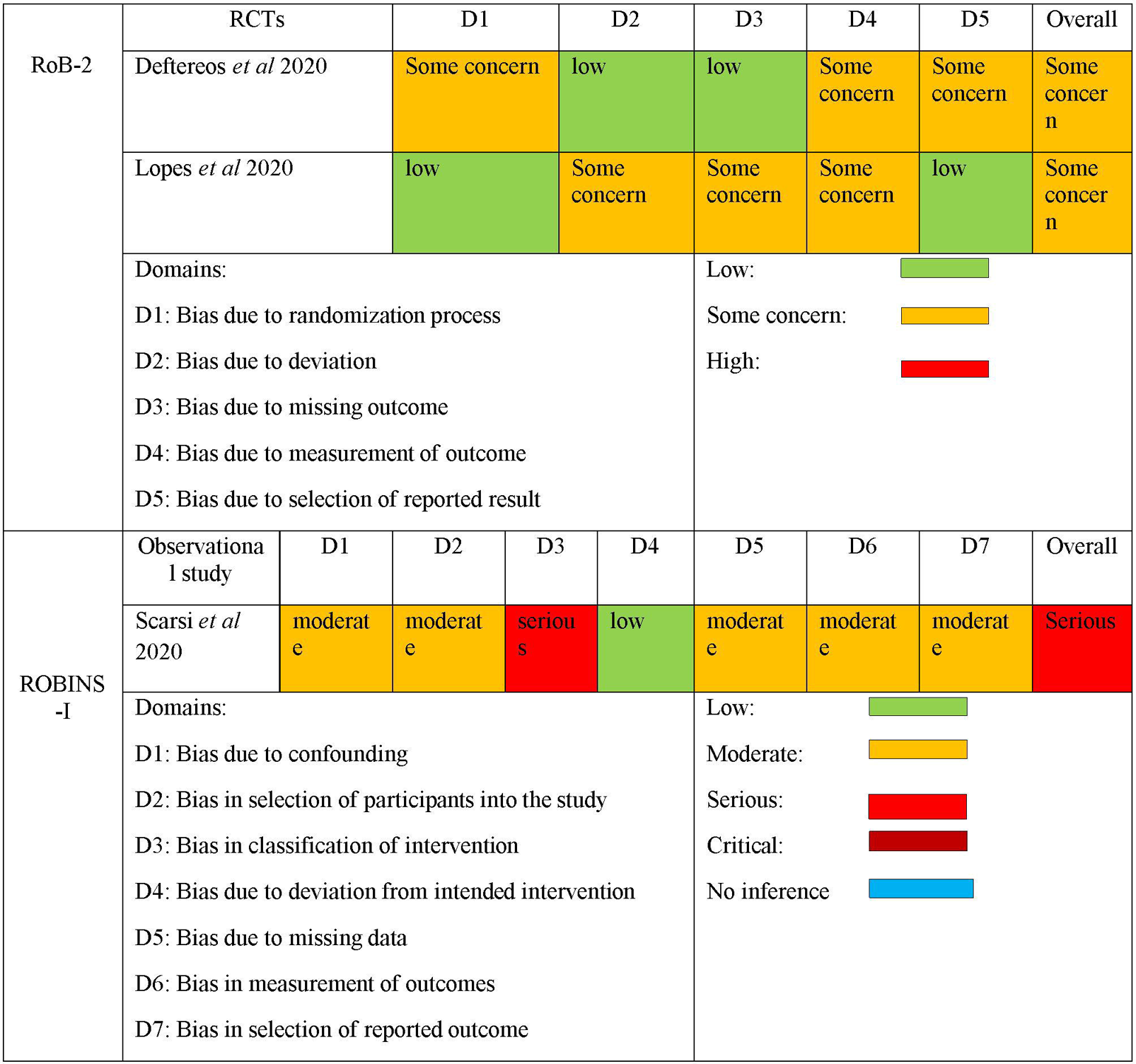
Risk of bias assessment

### Effects of intervention

The pooled effect of adding colchicine to usual therapy on primary and secondary outcomes was measured. The forest plots for the pooled effect are depicted in figure 3.

**Figure 3:**
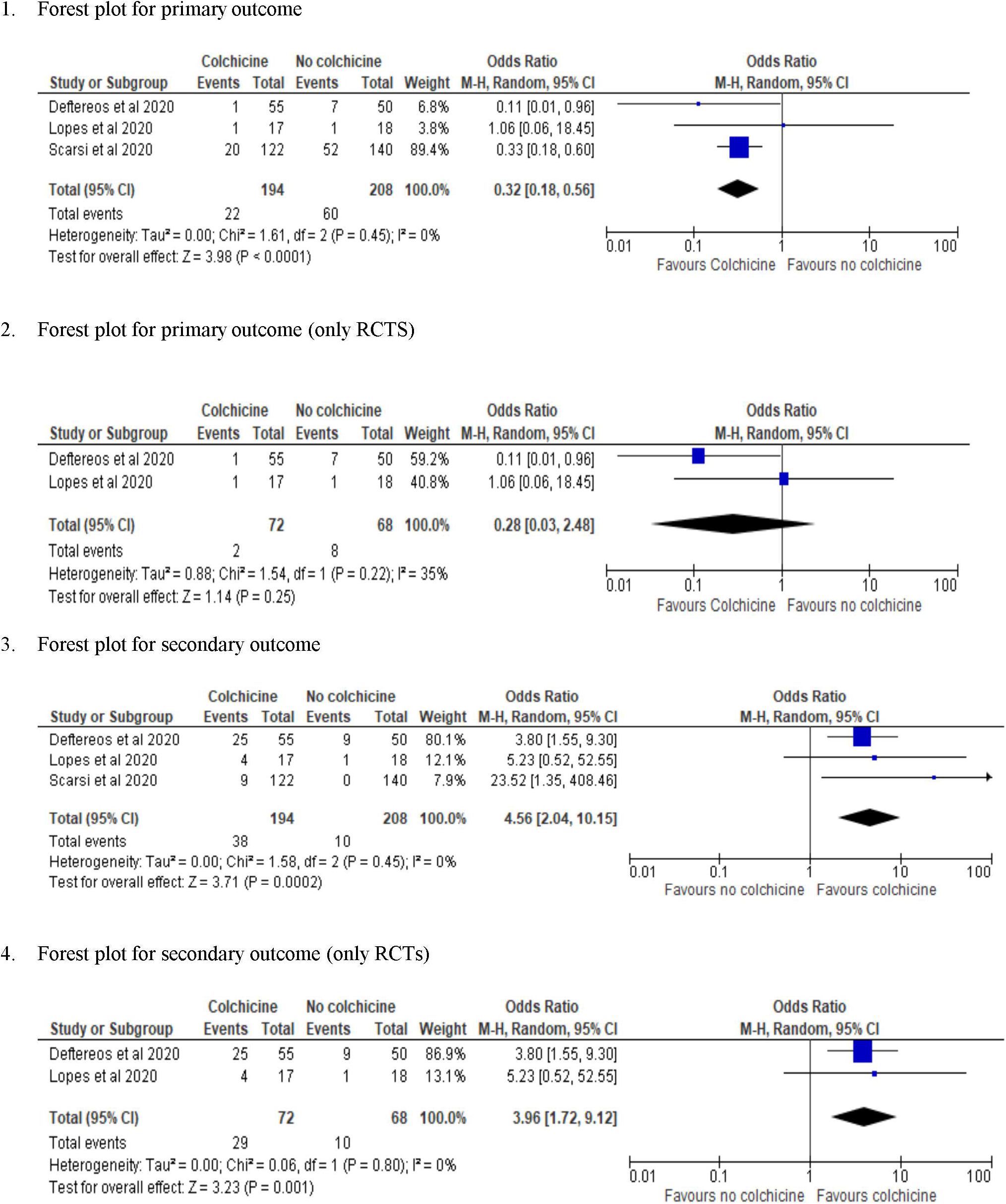
Forest plot

### Primary outcome: (Clinical deterioration)

Clinical deterioration was defined as the need for oxygen supplementation or ICU care or death within the follow-up period available in each study. The minimum and maximum duration of the follow-up period was 7 days and 21 days respectively. There was reduced reported clinical deterioration in all the included studies. In the study by Deftereos *et al*, 1 patient died in the colchicine group compared to 6 deaths and 1 ICU admission in the usual care group within a follow-up period of 21 days. However, Lopes *et al*, reported that 1 patient in each group required intensive care unit (ICU) admission with no death within a period of 7 days follow-up. In the study by Scarsi *et al*, there were 20 and 52 reported deaths in colchicine and usual care group respectively over a 21 days period. The overall pooled OR by random effect model was 0.32 (95%CI: 0.18 to 0.56) suggesting a statistically significant reduction in clinical deterioration with the addition of colchicine compared to usual care only (p <0.0001). Sensitivity analysis was not carried out as the test for heterogeneity for the pooled studies was not significant (Chi^2^ =1.61, df=2, (P=0.45), I^2^=0%). But subgroup analysis with only RCTs showed a reduction in clinical deterioration which was statistically non-significant (p= 0.25), pooled OR 0.28 (95% CI: 0.03 to 2.48).

### Secondary outcome

(Development of adverse effect) All the 3 included studies in the meta-analysis reported an increased incidence of adverse effects in the form of diarrhoea with colchicine use. The test of heterogeneity was not significant (Chi^2^=1.58, df=2, (P=0.45), I^2^ =0%). The random effect model revealed that there was a statistically significant increase in the adverse effects with colchicine use compared to usual therapy (OR=4.56, 95%CI: 2.04 to 10.15, P=0.0002). Sub-group analysis with only RCTs also revealed a statistically significant increase in the incidence of adverse effects with the use of colchicine (OR=3.96, 95%CI: 1.72 to 9.12, P=0.001).

### Publication bias

Although we created a Funnel plot for publication bias, it could not be assessed as there were only 3 studies where the primary outcome was reported. However, due to the small sample size and large effect we expect that publication bias would be present.

### Grade of evidence

The evidence was assessed using GRADE profiler for clinical deterioration (primary outcome) and the development of adverse effects (secondary outcome). The grade of evidence was low for both the outcomes due to the risk of bias in the included studies and publication bias.

The detailed analysis of the summary of evidence is depicted in table 3.

## Discussion

To the best of our knowledge, this is the first systematic review and meta-analysis on the use of colchicine in the management of COVID-19. In the current study, 402 patients from 3 countries were included for estimating the pooled effect. There was a statistically significant reduction in clinical deterioration with the addition of colchicine to usual therapy as suggested by the pooled estimate (p< 0.0001), though the effect became statistically non-significant when data from only the RCTs were analysed. The baseline characteristics, disease severity, and concomitant drug uses were comparable within both groups in all the studies. Scarsi *et al* reported that a lower risk of death was independently associated with colchicine treatment (HR=0.151 (95% CI 0.062 to 0.368), p<0.0001). The need for oxygen supplementation and duration of hospital stay was also lower in the colchicine group as reported by Lopes *et al*. However, RCTs are methodologically superior due to their design and their findings have higher validity than observational studies.

The observed adverse effects of colchicine use were similar across all the studies. Diarrhoea was the major adverse effect in all the included studies and was not severe in intensity. Other self-limiting adverse effects across the studies were nausea, vomiting, abdominal pain, headache, muscle cramps, and transient rise in liver enzymes in variable proportion. The pooled estimate suggested a statistically significant increase in the incidence of adverse effects (diarrhoea) with add on colchicine compared to usual care [OR=4.56, (P=0.0002)]. Diarrhoea was self-limiting in all the studies without any major electrolyte disturbance or dehydration. There was no need to stop the colchicine therapy in all included studies except for 2 patients on day 3 and day 6 as reported by Deftereos *et al*. These observations suggest that though the occurrence of adverse effect was statistically significant, it was not clinically important.

The role of colchicine in reducing D-dimer and inflammatory cytokine IL 6 were reported in many studies [16,21]. The attenuation of the D-dimer rise was also reported in the GRECCO-19 trial [18]. It is worth mentioning that the rise in D-dimer plays a major role in developing various complications in COVID-19. It is well documented in previous studies that colchicine exhibit anti-inflammatory action without immunosuppression unlike corticosteroids and IL-6 inhibitors [19]. This action might be beneficial for a certain subset of COVID-19 patients with primary or secondary immunosuppression.

### Limitations

The study has certain limitations. The number of included studies and the pooled sample size is less. Among them, one study is observational in nature and carries the risk of confounding and other biases. One of the RCTs included was open label and may have inherent assessment bias. The overall risk of bias in all studies ranged from some concern to serious and the quality of evidence is low. Additionally, there was variability in the follow up period between the studies ranging from 7 to 21 days.

## Conclusion

Colchicine is a safe, cost-effective, and potent anti-inflammatory medication without associated immunosuppression. Based on the limited number of available studies, the current meta-analysis shows a statistically significant reduction in clinical deterioration in COVID-19 patients with add on colchicine. Though there was an increase in adverse effects in the form of diarrhoea, it was mild and self-limiting. The clinical significance of repurposing colchicine for the management of COVID-19 will only emerge after larger RCTs with a greater sample size.

## Supporting information

Table 1: Characteristics of included studies

Table 2: Characteristics of patients included in the meta analysis

Table 3: Grade of evidence

## Data Availability

The data set has been available in pubMed AND medRxiv

## Author contribution

The development of the concept and the preliminary search was carried out by RRM and BMP. Extensive database search, data extraction, and quality assessment were carried out by RRM, SM, and BRM. Any disagreement was resolved by BMP. All the data synthesis and statistical analysis were done by BRM, BMP, and RRM. The manuscript was written by RRM, BRM, BMP, and SD. The final version for publication was approved by all the authors.

