## Supplementary material for "Repurposing Colchicine for the Management of COVID-19: A Systematic Review and Meta-analysis": Table 1: Characteristics of included studies

| Study name | Deftereos *et al* 2020  3^rd^ April to 27^th^ April 2020 | Lopes *et al* 2020  11^th^ April to 6^th^ July 2020 | Scarsi *et al* 2020  5^th^ March to 5^th^ April 2020 |
| --- | --- | --- | --- |
| Country | Greece | Brazil | Italy |
| Type of study | Prospective, open-label, randomized clinical trial | Randomized, double-blinded, placebo controlled clinical trial | Prospective cohort study |
| Sample size | 105 | 35 | 262 |
| Inclusion criteria | Age >18 years, both gender  Symptomatic & RT-PCR confirmed COVID-19  QT interval <450 millisecond. | Age >18 years, both gender  RT-PCR confirmed moderate to severe COVID-19 pneumonia  QT interval <450 millisecond. | Virologicaly and radiographically confirmed COVID-19 of both gender |
| Primary outcome | Biochemical phase- maximum high sensitivity cardiac troponin level, time for C- reactive protein to reach more than 3 times of upper limit of reference  Clinical phase- time to deterioration by 2 points on a 7-point clinical status scale | Clinical parameters ranging from requirement of supplemental oxygen to death | All-cause mortality |
| Secondary outcome | All-cause mortality  Requirement of mechanical ventilator  Adverse effects | Change in clinical parameter  Adverse effects | Adverse effects |
| Follow up period | 21 days | 7 days | 21 days |
| Colchicine dose | A loading dose of 1.5 milligram (mg) followed by 0.5 mg after 1 hour. Maintenance dose of 0.5 mg twice daily | 0.5 mg 3 times daily for 5 days followed by 0.5 mg 2 times daily for 5 days. | 1mg per day |
