## Supplementary material for "Repurposing Colchicine for the Management of COVID-19: A Systematic Review and Meta-analysis": Table 2: Characteristics of patients included in the meta analysis

|  | Deftereos *et al* 2020 | | Lopes *et al* 2020 | | Scarsi *et al* 2020 | |
| --- | --- | --- | --- | --- | --- | --- |
|  | Colchicine | No colchicine | Colchicine | No colchicine | colchicine | No colchicine |
| Patient characteristics | n=55  Male- 31  Female- 24  Median age (IQR)- 63 (55 to 70) | n=50  Male- 30  Female- 20  Median age (IQR)- 65 954 to 80) | n= 17  Male- 9  Female- 8  Median age (IQR)-  48 (41.5 to 64) | n= 18  Male- 5  Female- 13  Median age (IQR)-  53.5 (35.5 to 65.5) | n=122  Male- 77  Female- 45  Mean age (SD)-  69.3 (9.6) | n=140  Male- 90  Female- 50  Mean age (SD)-  70.5 (13.8) |
| Clinical deterioration (Primary outcome) | 1 | 7 | 1 | 1 | 20 | 52 |
| Development of adverse effect (Secondary outcome) | 25 | 9 | 4 | 1 | 9 | 0 |
| Comorbidities  [ n (%)] | Hypertension 22 (40)  Diabetes  9 (16)  Coronary artery disease  9 (16)  Respiratory disease  3 (5) | Hypertension 25 (50)  Diabetes  12 (24)  Coronary artery disease  5 (10)  Respiratory disease  2 (4) | Diabetes  5 (29)  Coronary artery disease  8 (47)  Respiratory disease  3 (18) | Diabetes  6(33)  Coronary artery disease  6 (33)  Respiratory disease  2 (11) | Coronary artery disease  65(64)  Respiratory disease  17(17) | Coronary artery disease  85(74)  Respiratory disease  24(22) |
| Additional treatment received | Hydroxychloroquine, Azithromycin, Lpinavir, Tocilizumab, Anticoagulant | Hydroxychloroquine, Azithromycin, Lpinavir, Tocilizumab, Anticoagulant | Hydroxychloroquine, Azithromycin, Heparin, Steroid | Hydroxychloroquine, Azithromycin  Heparin, Steroid | Hydroxychlorquine, Lopinavir, Steroid | Hydroxychlorquine, Lopinavir, Steroid |
