## Supplementary material for "Repurposing Colchicine for the Management of COVID-19: A Systematic Review and Meta-analysis": Table 3: Grade of evidence

| **Question: Should Colchicine be used for COVID-19? Bibliography:** Meta-analysis | | | | | | | | | | | |
| --- | --- | --- | --- | --- | --- | --- | --- | --- | --- | --- | --- |
| **Quality assessment** | | | | | | | **Summary of Findings** | | | | |
| **Participants (studies) Follow up** | **Risk of bias** | **Inconsistency** | **Indirectness** | **Imprecision** | **Publication bias** | **Overall quality of evidence** | **Study event rates (%)** | | **Relative effect** (95% CI) | **Anticipated absolute effects** | |
|  |  |  |  |  |  |  | **With Control** | **With Colchicine** |  | **Risk with Control** | **Risk difference with Colchicine** (95% CI) |
| **Clinical deterioration** (assessed with: Need for oxygen supplementation/ ICU care/ death) | | | | | | | | | | | |
| 402 (3 studies^1^) | serious | no serious inconsistency | no serious indirectness | no serious imprecision | reporting bias strongly suspected | ⊕⊕⊝⊝ **LOW** due to risk of bias, publication bias | 60/208  (28.8%) | 22/194  (11.3%) | **OR 0.31**  (0.18 to 0.54) | **Study population** | |
|  |  |  |  |  |  |  |  |  |  | **288 per 1000** | **177 fewer per 1000** (from 109 fewer to 220 fewer) |
|  |  |  |  |  |  |  |  |  |  | **Moderate** | |
|  |  |  |  |  |  |  |  |  |  |  | **-** |

| **Development of adverse effect** (assessed with: Development of diarrhea (as it is the most common side effect with colchicine) and/or any other side effect leading to withdrawal of colchicine. ) | | | | | | | | | | | |
| --- | --- | --- | --- | --- | --- | --- | --- | --- | --- | --- | --- |
| 402 (3 studies^1^) | serious | no serious inconsistency | no serious indirectness | no serious imprecision | reporting bias strongly suspected | ⊕⊕⊝⊝ **LOW** due to risk of bias, publication bias | 10/208  (4.8%) | 38/194  (19.6%) | **OR 5.31**  (2.43 to 11.59) | **Study population** | |
|  |  |  |  |  |  |  |  |  |  | **48 per 1000** | **163 more per 1000** (from 61 more to 321 more) |
|  |  |  |  |  |  |  |  |  |  | **Moderate** | |
|  |  |  |  |  |  |  |  |  |  |  | **-** |

^1^ One observational study
